# User involvement in the design and development of medical devices in Epilepsy: a systematic review

**DOI:** 10.1101/2024.07.24.24310932

**Authors:** João Ferreira, Ricardo Peixoto, Lígia Lopes, Sándor Beniczky, Philippe Ryvlin, Carlos Conde, João Claro

**Author notes:** Authors for correspondence: João Ferreira ORCID: 0000-0002-5827-0623. Rua Sousa Aroso 31 Ed.2 1°B 4450-289 Matosinhos, Portugal; Carlos Conde ORCID: 0000-0002-4177-8519. Rua Alfredo Allen 208, 4200-135, Porto, Portugal; João Claro ORCID: 0000-0001-5936-1036 INESC TEC and Faculdade de Engenharia, Universidade do Porto, Campus da FEUP, Rua Dr. Roberto Frias, 4200-465 Porto, Portugal.

## Abstract

**Objective:** This systematic review aims to describe the involvement of persons with epilepsy (PWE), healthcare professionals (HP) and caregivers (CG) in the design and development of medical devices is epilepsy.

**Methods:** A systematic review was conducted, adhering to the Preferred Reporting Items for Systematic Reviews and Meta-Analyses (PRISMA) guidelines. Eligibility criteria included peer-reviewed research focusing on medical devices for epilepsy management, involving users (PWE, CG, and HP) during the MDD process. Searches were performed on PubMed, Web of Science, and Scopus, and a total of 55 relevant articles were identified and reviewed.

**Results:** From 1999 to 2023, there was a gradual increase in the number of publications related to user involvement in epilepsy medical device development (MDD), highlighting the growing interest in this field. The medical devices involved in these studies encompassed a range of seizure detection tools, healthcare information systems, vagus nerve stimulation (VNS) and electroencephalogram (EEG) technologies reflecting the emphasis on seizure detection, prediction, and prevention. PWE and CG were the primary users involved, underscoring the importance of their perspectives. Surveys, usability testing, interviews, and focus groups were the methods employed for capturing user perspectives. User involvement occurs in four out of the five stages of MDD, with production being the exception.

**Significance:** User involvement in the MDD process for epilepsy management is an emerging area of interest holding a significant promise for improving device quality and patient outcomes. This review highlights the need for broader and more effective user involvement, as it currently lags in the development of commercially available medical devices for epilepsy management. Future research should explore the benefits and barriers of user involvement to enhance medical device technologies for epilepsy.

**Plain Language Summary:** This review covers studies that have involved users in the development process of medical devices for epilepsy. The studies reported here have focused on getting input from people with epilepsy, their caregivers, and healthcare providers. These devices include tools for detecting seizures, stimulating nerves, and tracking brain activity. Most user feedback was gathered through surveys, usability tests, interviews, and focus groups. Users were involved in nearly every stage of device development except production. The review highlights that involving users can improve device quality and patient outcomes, but more effective involvement is needed in commercial device development. Future research should focus on the benefits and challenges of user involvement.

**Key Point Box:**

- PWE are the users more involved in the MDD process;
- Surveys and usability testing are the methods more frequently used for user involvement in the MDD process in epilepsy;
- Literature only discloses the involvement of users in the MDD process of 13 commercially available medical devices for epilepsy management.

## 1. INTRODUCTION

Epilepsy impacts approximately 70 million individuals and 30 % of them do not respond to current treatments to control their seizures ^1^. Accurately anticipating the onset of seizures and managing them can assist PWE in avoiding self-injury and potentially enhance their overall well-being. Seizure detection and, more recently, seizure forecasting are critical areas of clinical advancement in epilepsy. The progress in these areas has been driven by developments in medical devices, which have the potential to optimize seizure control and prevent seizure-related morbidity and mortality in individuals with epilepsy ^2^.

Medical devices used in epilepsy range from implantable devices such as vagus nerve stimulators, responsive neurostimulation and deep brain stimulation to non-implantable devices such as electroencephalography (EEG) based systems and non-EEG based seizure detection wearable devices ^1^. Regardless of the type of medical device, the ultimate goal is to develop devices that meet the needs of end-users (PWE, CGs and HP), improve disease outcomes, and enhance the overall quality of life of individuals with epilepsy.

Medical device manufacturers create life-changing innovations through the collaborative expertise of various disciplines, including engineering, manufacturing, clinical, regulatory, marketing, sales, and business specialists. While cutting-edge technology advancement in medical device design is absolutely vital, it is the overall experience (cognitive and emotional) that impacts the daily life of the patient and CG ^3^. In recent years, there has been a growing recognition of the importance of user involvement in MDD in general ^4^, and specifically for epilepsy ^5^. In epilepsy, user involvement refers to the active participation of PWE, CGs, HP and other stakeholders in the design and development of medical devices. Understanding and incorporating users’ needs, preferences, and feedback into the MDD process can help to ensure that the device is effective, safe, and well-received by the intended users. User involvement can take various forms, including needs assessment, usability testing, co-creation and co-design ^6^. User-centred design is a critical factor in the design and development of medical devices. Specifically, considering user needs during the early stages of device conceptualization and throughout the subsequent development process can yield substantial benefits. This approach can improve patient safety, increase compliance with treatment regimens, and enhance health outcomes ^7^. Additionally, user-centred design promotes higher levels of user satisfaction, and it can lead to a reduction in device development time by identifying and addressing usability issues prior to launch. This, in turn, can help to avoid costly design changes and product recalls, which can have significant financial and reputational implications for device manufacturers ^8^. Therefore, successful medical device innovation requires investigation of end-user and broader stakeholder contexts and incorporation of those context-specific needs into design processes ^9^.

Despite the initial promise that user involvement in medical device design and development for epilepsy holds, there is widespread scepticism regarding its effectiveness within the scientific, medical, and general communities, leaving important choices and questions open for debate, namely: (i) who should be involved, (ii) at which stages should they be involved, (iii) which participatory methods are most suitable and (iv) what topics are to be discussed with end users during development. Therefore, we conducted a systematic review to explore the practice of user involvement in the design and development of medical devices for epilepsy, aiming to provide valuable insights into the role of user involvement in this context and inform future research and practice in this area. Descriptive statistics and qualitative thematic analysis were used for analysing the data, which were divided into different themes, i.e., types of medical devices developed and assessed; types of medical device users involved; extent of user involvement by different stages of the MDD cycle; and methods used for capturing users’ perspectives.

## 2. METHODS

The present systematic literature review was performed to identify and extract all currently available literature related to user involvement in medical devices or technology utilized in the monitoring, treatment and/or management of epilepsy.

### 2.1 Protocol and registration

This systematic review was reported according to the Preferred Reporting Items for Systematic Reviews and Meta-Analyses (PRISMA) guidelines ^10^, and was prospectively registered on the International Prospective Register of Systematic Reviews (PROSPERO) (CRD42023490599).

### 2.2 Eligibility Criteria

Publications inclusion and eligibility for short-listing criteria encompassed (a) peer-reviewed original research, (b) targeted only at medical devices according to WHO’s definition ^11^ with application for epilepsy management and (c) indicating user (PWE, CG and HP) involvement during these products’ development lifecycle.

Original publications were excluded if presenting any of the following characteristics: the wrong population (i.e., non-human population); the wrong intervention (i.e., medical devices used for other purposes rather than epilepsy management); the wrong outcome (i.e., there were no epilepsy or seizure diagnosis, management, or treatment outcomes); publication not available in the English language; publications not in full publication. All studies published up to 30 November 2023 were included with no other time limitations.

### 2.3 Search strategy

Search strings related to medical devices, users’ (PWE, CG and HP) involvement and epilepsy were developed. Thus, the search term (“epilepsy”) was searched in combination with the following search strings: (“device users” OR “end-users” OR “medical devices” OR “medical device users” OR “needs assessment” OR “new medical technology” OR “user centered product” OR “user criteria” OR “user input” OR “user interests” OR “user involvement” OR “user needs” OR “user needs assessment” OR “user needs research” OR “user participation” OR “user perceptions” OR “user perspective” OR “user requirements” OR “user requirements elicitation” OR “user studies” or “user survey” OR “user-based” OR “wearable device” OR “device” and “acceptability” OR (“device” AND “usability”) OR (“device” AND “satisfaction”) OR (“device” AND “patient views”)). The Boolean operator AND was used to link the search term with the respective search strings on all databases.

Three databases were used from database inception on 30 November 2023: PubMed, Web of Science and Scopus. All searches were performed based on the title, abstract and keyword in all databases. Articles were first screened through their titles and abstracts before proceeding with the full-text screening of relevant articles. The search criteria and keywords were arrived at through consensus with all researchers. Hand searching, personal collections (unpublished studies, conference abstracts, grey literature, or other resources that researchers have accumulated through personal networks or professional contacts) and exploding references (reviewing the reference lists of relevant articles to identify additional studies that may not have been captured through our primary search strategy) were also used to augment the results.

### 2.4 Selection Process

Database search results were imported into EndNote (EndNote x9; Clarivate, Philadelphia, PA, USA), and duplicates were removed. Results were then exported to an Excel spreadsheet for title, abstract, and full-text screening. Study selection, data extraction and assessment of study quality followed the Standards for Reporting Qualitative Research^13^. No assumptions were made during the selection to reduce the risk of bias. Each paper was screened and discussed by two independent reviewers (JF, RP). Disagreements were resolved by a third independent researcher (CC).

### 2.5 Data collection and analysis

Data was extracted using a data-extraction form (Appendix 1.) developed for the review ^14^. Data extracted included: (1) author details, (2) year of publication, (3) country of study, (4) type of medical device, (5) method/tool/approach, (6) users and (7) product development stages. Since there is no standard framework to describe the MDD stages ^15^, for data extraction purposes, the medical device lifecycle was divided into five stages: 1- Concept; 2- Design; 3-Testing and Trials; 4- Production; 5-Deployment. The classification of medical device lifecycle stages was carried out based on the product lifecycle reported in the literature (Table 1 adapted from Shah, Robinson ^16^).

**Table 1.**
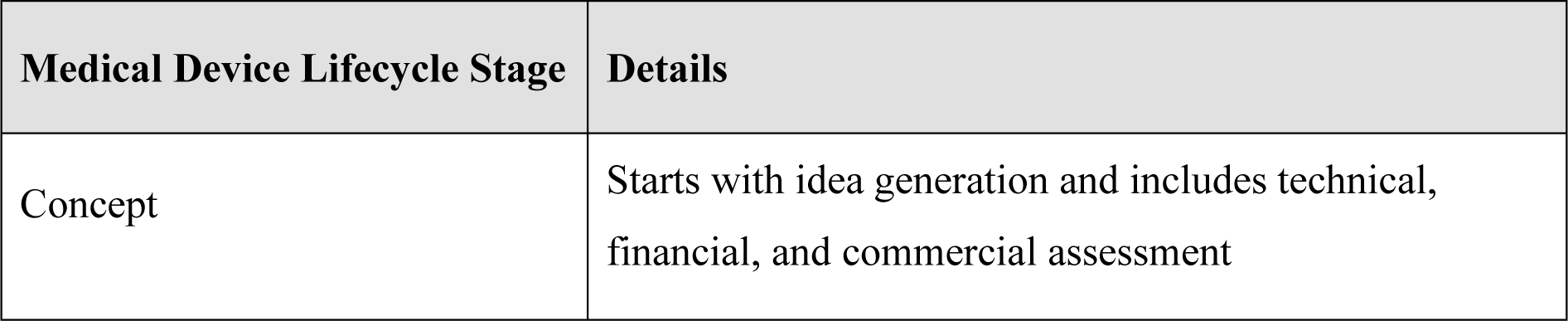
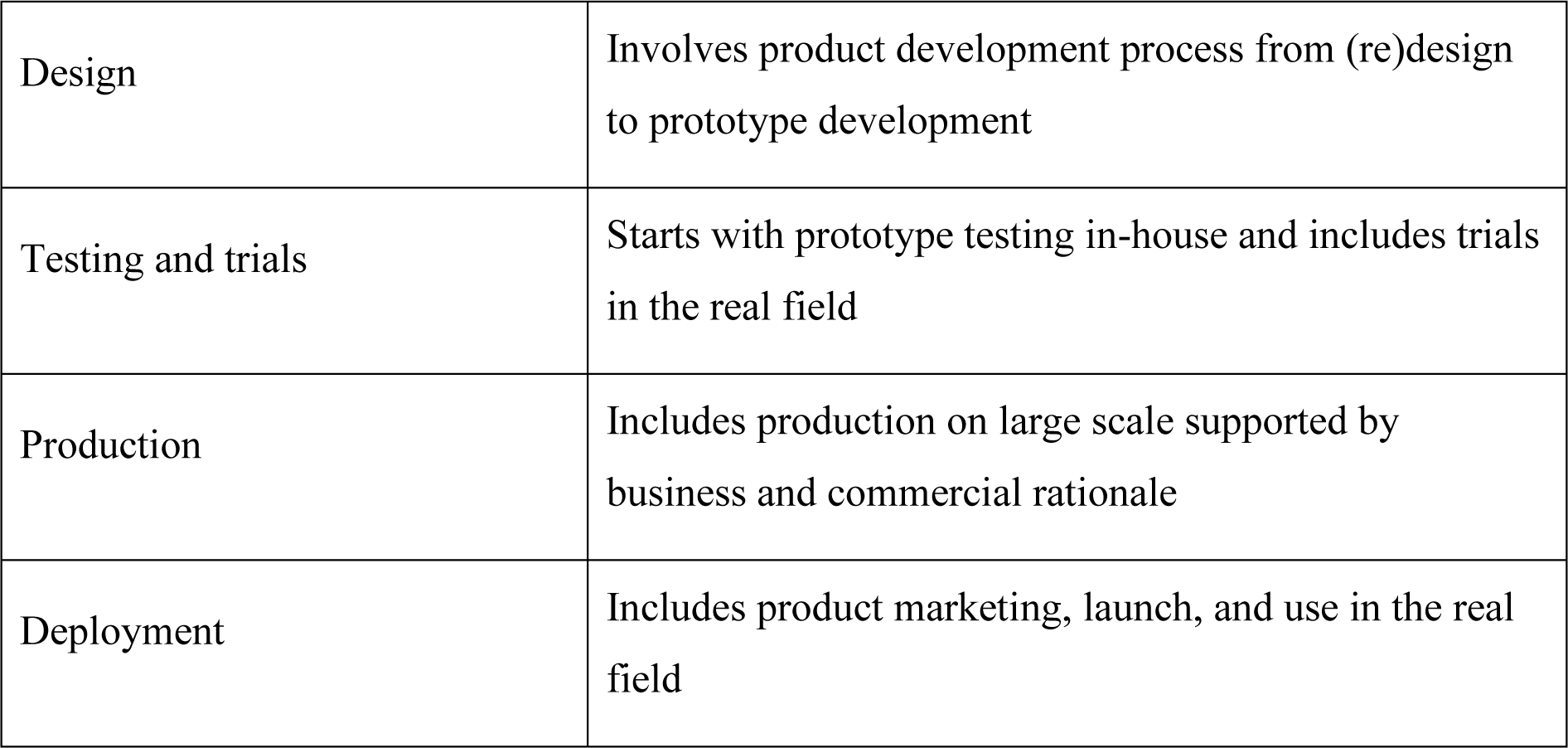
Product Lifecycle Stages (Adapted from Shah, Robinson ^16^)

| Medical Device Lifecycle Stage | Details |
| --- | --- |
| Concept | Starts with idea generation and includes technical, financial, and commercial assessment |
| Design | Involves product development process from (re)design to prototype development |
| Testing and trials | Starts with prototype testing in-house and includes trials in the real field |
| Production | Includes production on large scale supported by business and commercial rationale |
| Deployment | Includes product marketing, launch, and use in the real field |

## 3. RESULTS

The study workflow and paper selection process is illustrated in Figure 1. Following the inclusion criteria review, 55 publications (^17 18 19 20 21 22 23 24 25; 26; 27; 28; 29; 30; 31; 32; 33; 34; 35; 36; 37; 38; 39; 40; 41; 42; 43; 44; 45; 46; 47; 48; 49; 50; 51; 52; 53; 54; 55; 56; 57; 58; 59; 60; 61; 62; 63; 64; 65; 66; 67; 68; 69; 70; 71^) were included, and 109 were excluded (see Figure 1 for PRISMA flow chart).

**Figure 1.**
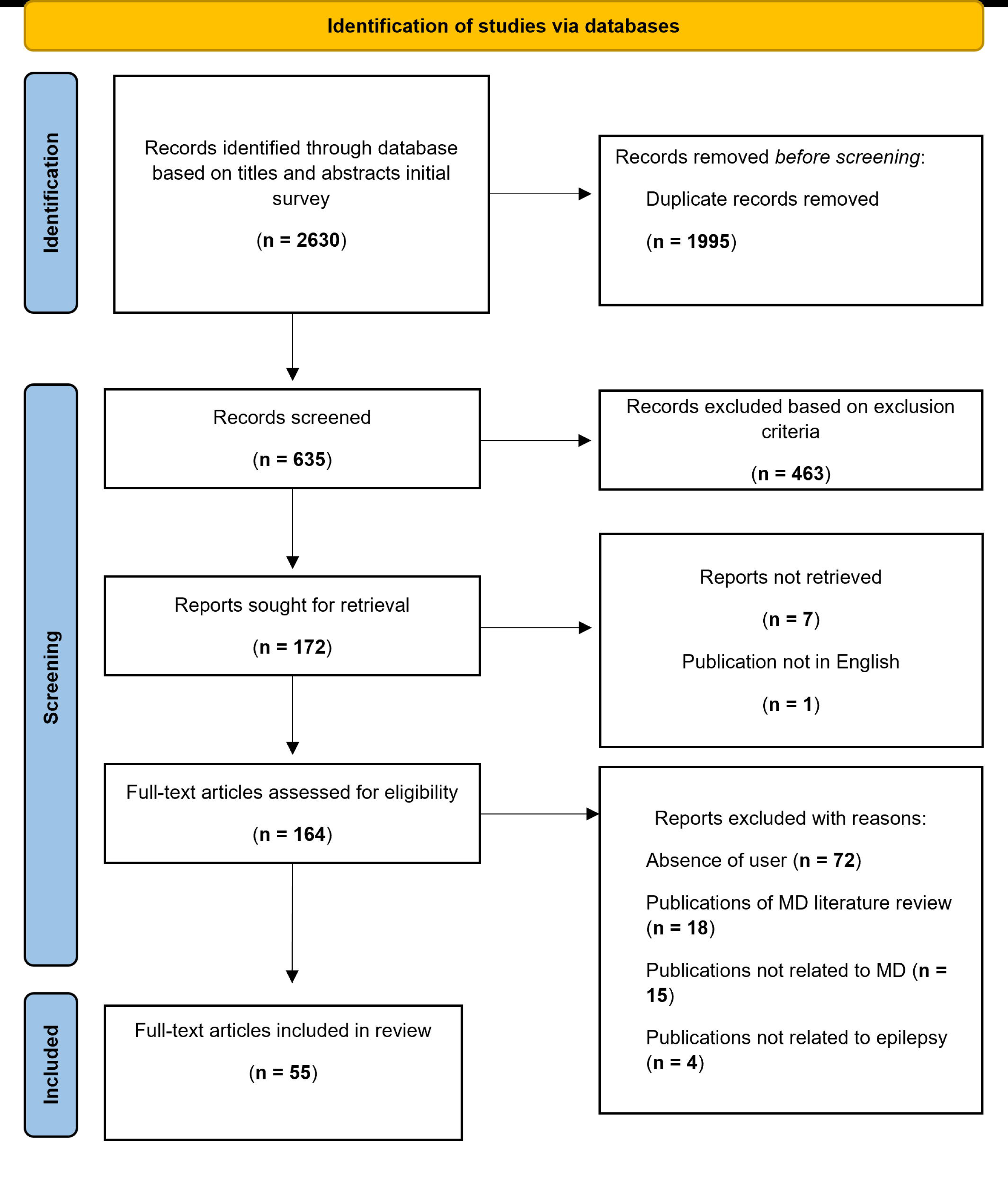
PRISMA Flowchart adopted for the present systematic review.

Seventeen of the 55 included publications were from the United States, 7 from the United Kingdom, 6 from Canada, 5 from Denmark, 4 from The Netherlands, 3 from Germany and 3 from South Korea and 1 from Sweden, 1 from Mexico, 1 from France, 1 from Italy and 1 from Belgium. Five studies were of multiple origin (Table S1).

Across the years, we find that user involvement in MDD is becoming more frequent. Figure 2 shows the number of papers published per year and the total accumulated number of publications over time since the inaugural paper by Hufford, R.L., P.M. ^72^.

**Figure 2.**
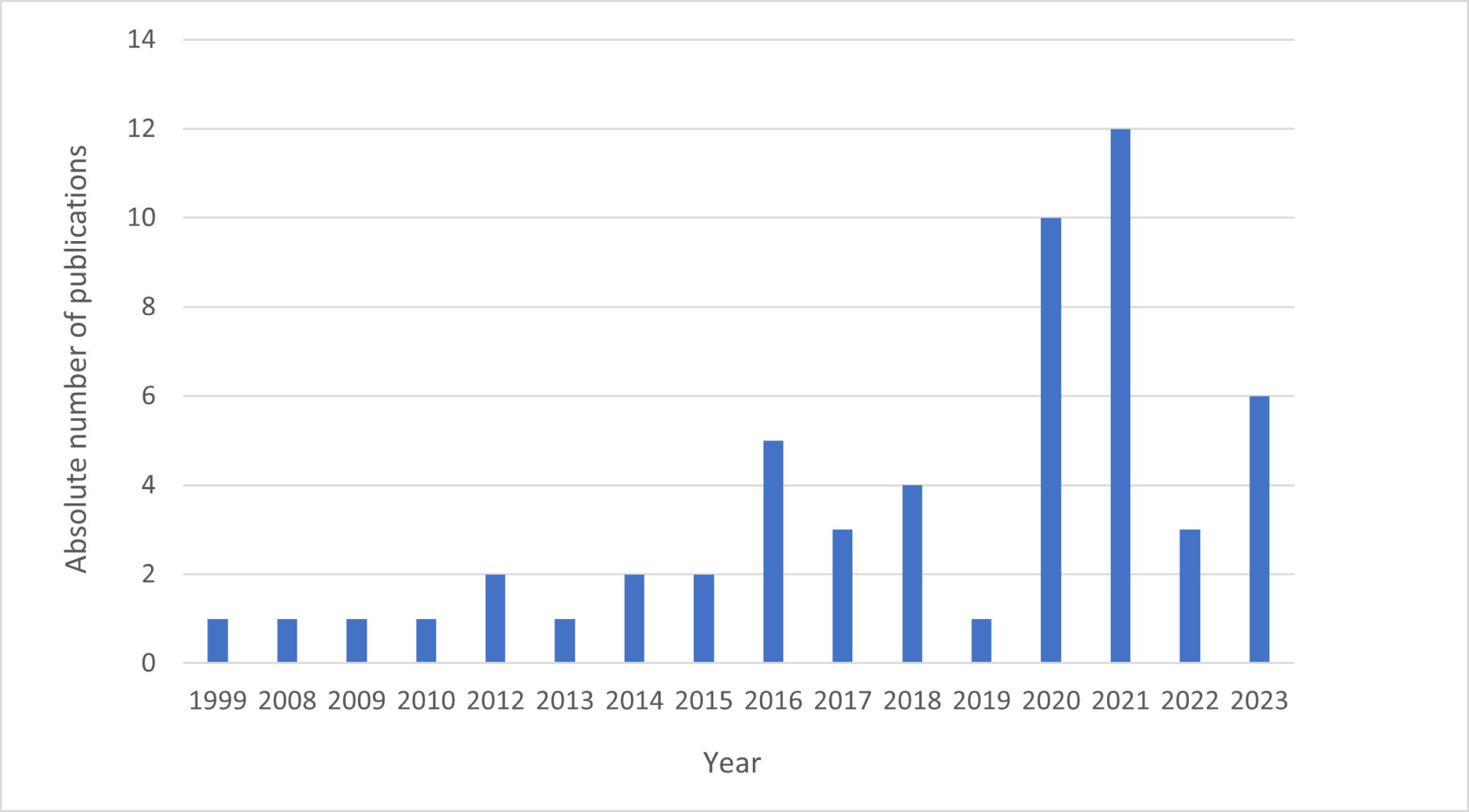
Number of publications per year (blue bars) and cumulative number (orange line) of publications from 1999 to 2023, as of *November 30, 2023. (N=55)

### 3.1 Types of medical devices assessed by user involvement

In our comprehensive review, we have identified a broad spectrum of medical devices (as illustrated in Figure 3) that have been conceived and rigorously assessed by involving end-users. The majority of the devices were seizure detection devices (44%) and healthcare / clinical information systems (40%) such as Software as Medical Device, web-based tools, electronic health records, and videoconferencing systems. Vagus Nerve Stimulator systems represented 9% of the studies found, followed by EEG (5%) and 1 study on an automated injection system (2%).

**Figure 3.**
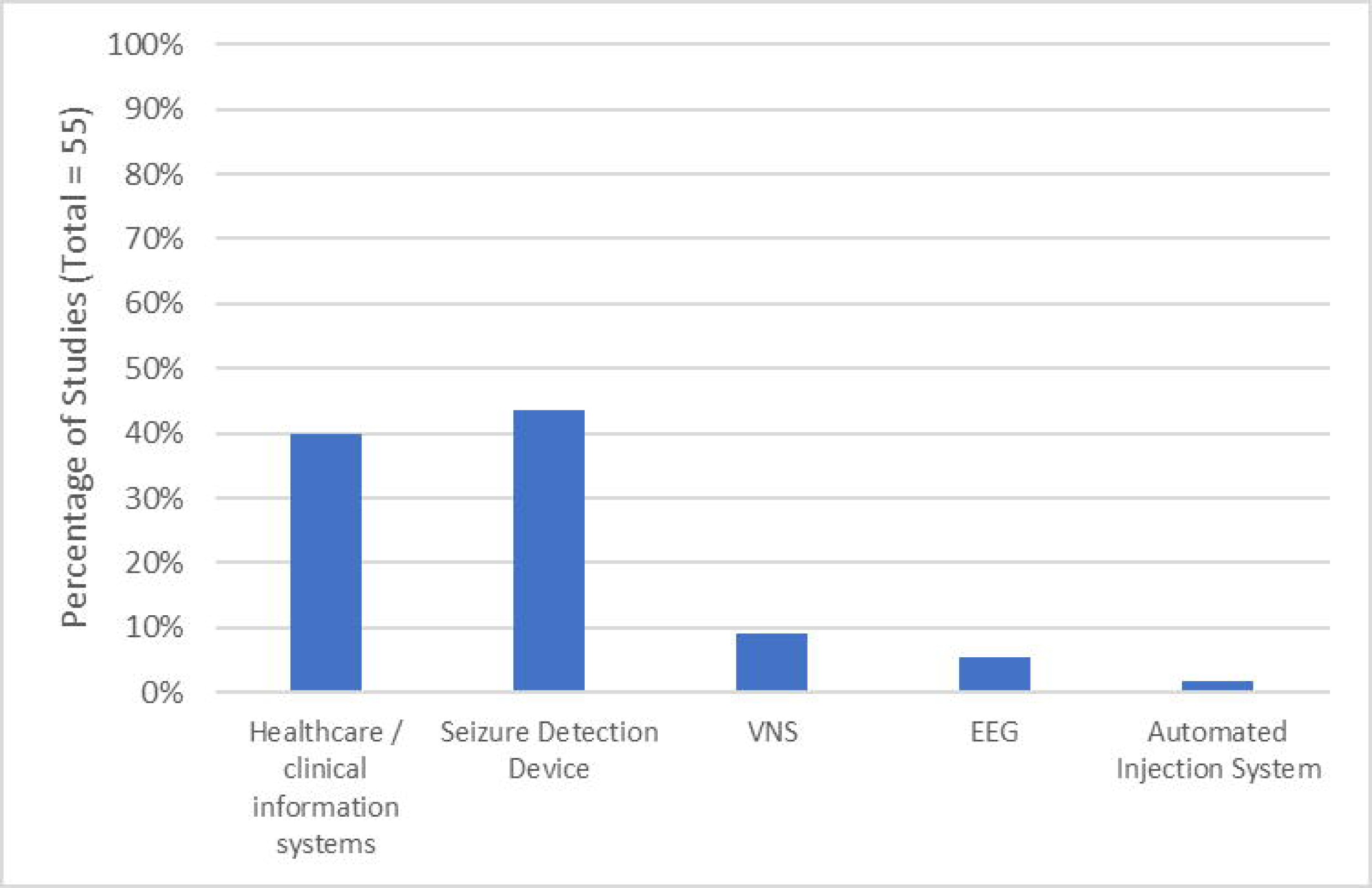
Types of medical devices developed and assessed by user involvement (N=55)

### 3.2 Types of medical devices users involved in MDD and assessment

A wide range of users were involved in the MDD process, including clinicians, PWE, carers, family members and persons with different disabilities and impairments (Figure 4). PWE were the users more frequently involved (40%, N = 3), followed by the PWE alongside CG (27%, N = 15) and HP (11%, N = 6). CG and HP alone were the users less frequently involved.

**Figure 4.**
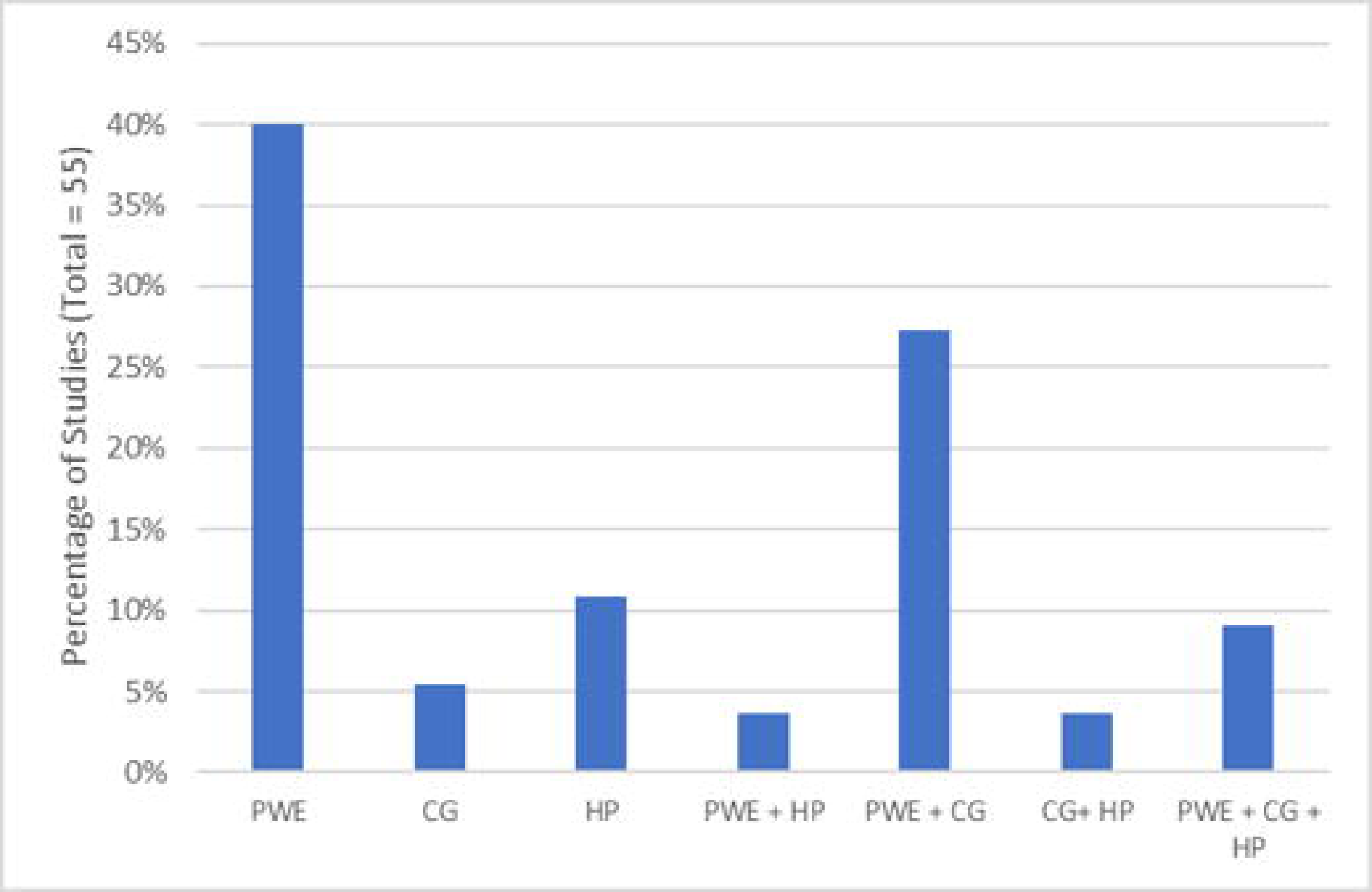
Included publications by types of users (N=55), CG – caregivers; HP – Health Professionals; PWE – Patients with Epilepsy.

### 3.3 Extent of user involvement by stage of the medical device lifecycle

The findings of the selected studies, especially the stages of the development of new medical devices, and methods used for capturing user perspectives by stages of the medical device lifecycle, are summarized in Figure 5 and Table S1. In 93% (N= 51) of the studies, users were involved in one stage of the medical device lifecycle especially in the deployment stage, and in the remaining studies users were involved in 2 (5%, N = 3) or 3 (2%, N = 1) stages of the lifecycle. Single-stage involvement was predominant in the deployment phase during which the product is already in the market, as reported in 45% (N = 25) of the studies, followed by concept stages in 25% (N = 14), when all uncertainties such as the clinical need definition, customer requirements and needs, finances, reimbursement strategy, team selection, or legal aspects must be considered. Two-stage combinations were between design and testing and trials in 5% (N = 3). Three-stage combinations for user involvement were between concept, design, and testing and trials stages in 2% (N = 1), which is shown in Figure 5.

**Figure 5.**
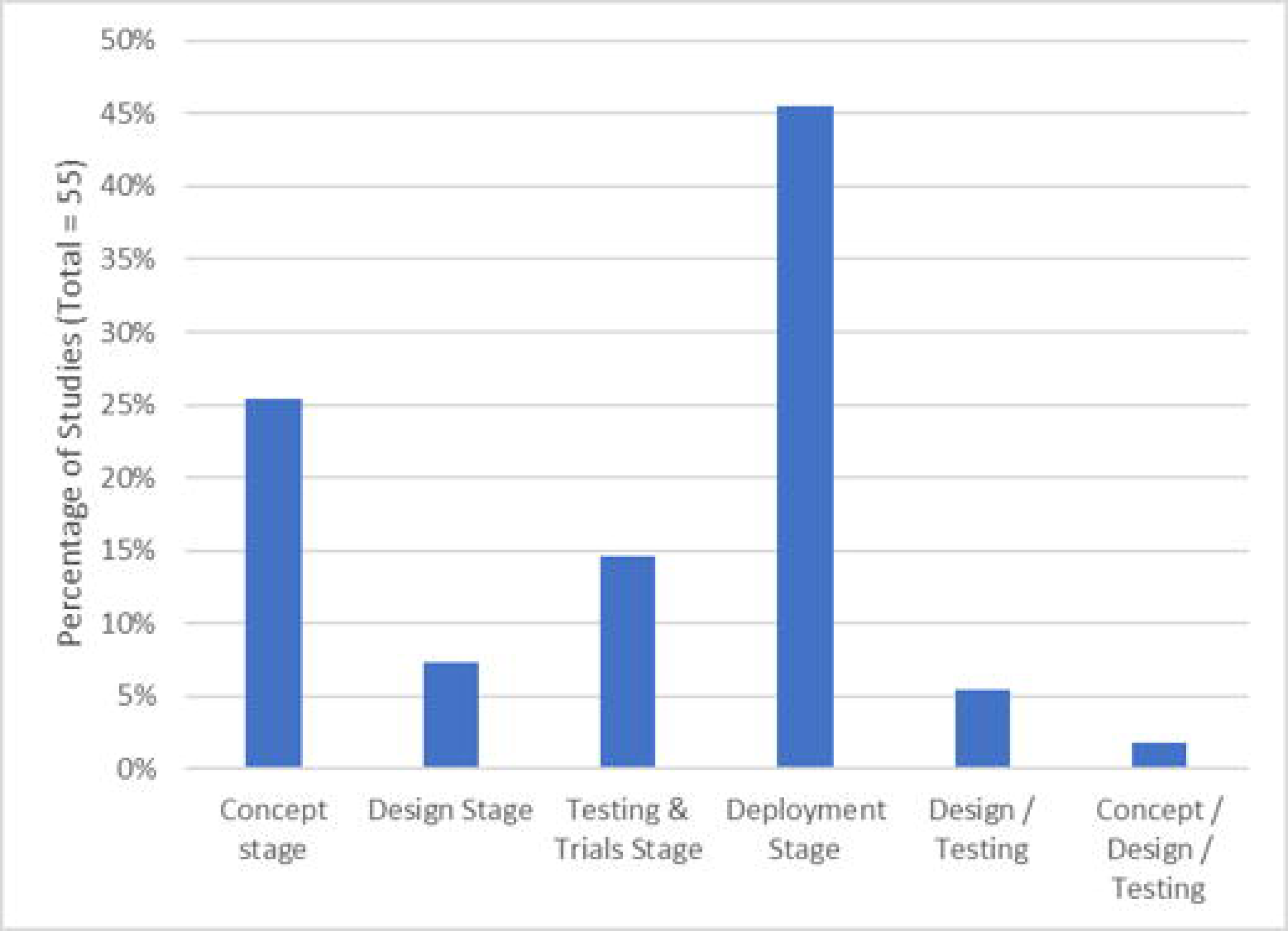
Included publications by products lifecycle stages (N=55)

All other studies are less comprehensive as their models only encompass one stage.

### 3.4 Methods used for capturing users’ perspectives

Our review shows that several methods were designed to involve users, often requiring the combination of both qualitative methods to evaluate parameters from a numerical point of view and quantitative methods to indicate the user’s choices, thoughts, and feelings. Involving users and capturing their perspectives in the medical device technology lifecycle concerned mostly surveys in 49% (N=43), usability testing in 20% (N=18), interviews in 19% (N=17), and focus groups in 11% (N=10) of the studies (Figure 6). These methods are mapped against the medical device lifecycle stages where they were used (Table 2). Some of the studies used more than one method of inquiry.

**Figure 6.**
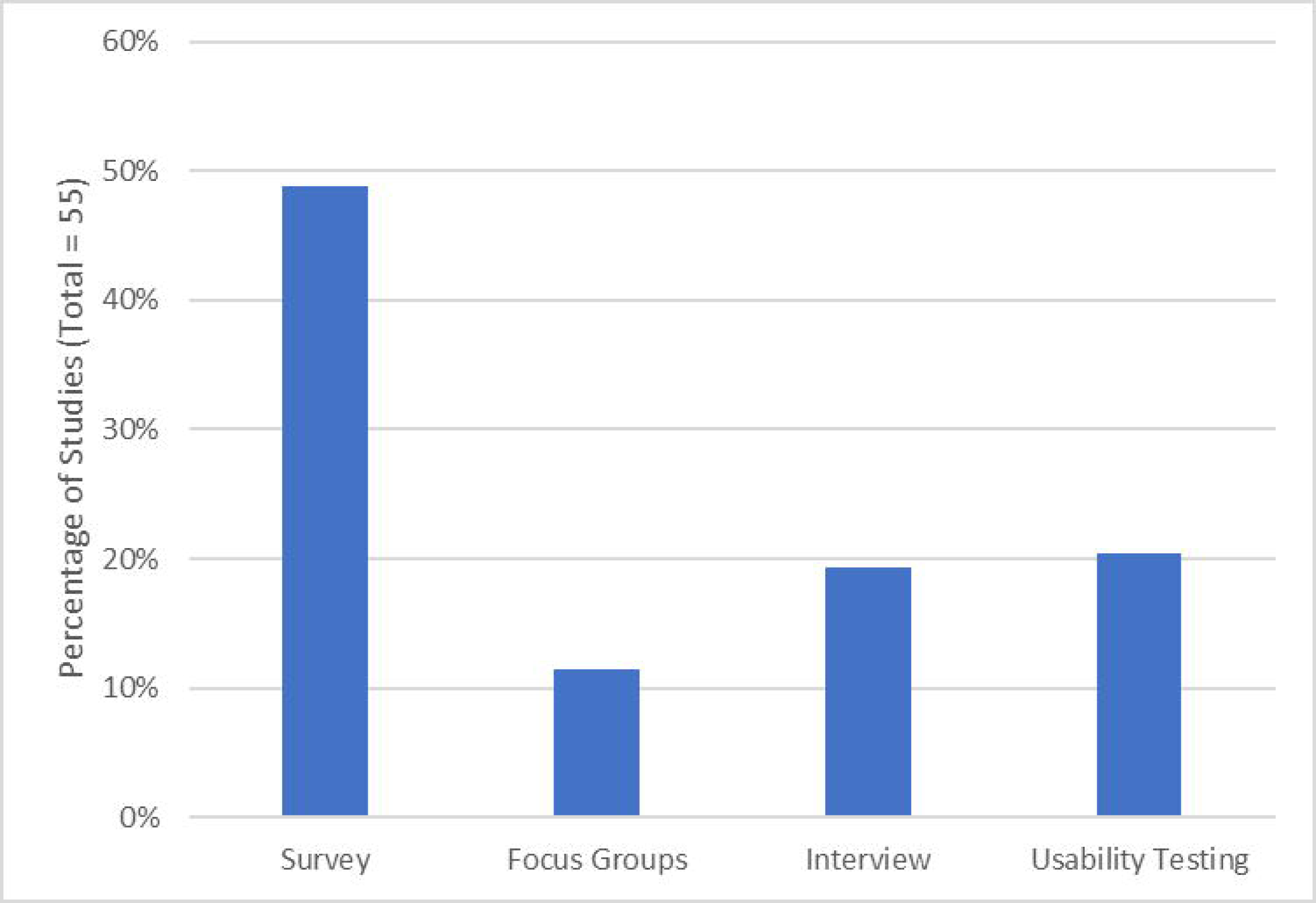
Methods used for capturing user perspectives (N=55).

**Table 2.** Methods used for capturing user perspectives by stages of the medical device lifecycle (N=55)

| Concept Stage | Design Stage | Test and trials stages | Deployment stage |
| --- | --- | --- | --- |
| Survey (11) | Focus Groups (4) | Usability Testing (6) | Interviews (10) |
| Focus Groups (2) | Interviews (3) | Survey (8) | Survey (16) |
| Interview (3) | Survey (4) | Focus Groups (3) | Usability Testing (6) |
| Usability Testing (1) | Usability Testing (4) |  | Focus Groups (3) |

*N – total publications included*

Surveys and focus groups were used in all four medical device lifecycle stages where the users were involved. Typically, surveyed individuals were asked to respond to the questions in a yes/no manner, on a Likert-type scale (e.g., very often to not at all often), or with open-ended responses. The choice of responses was dictated by the investigator and the medical device (if one was used). The selection of the type of response desired was often made based on the difficulty of the question asked and the depth of knowledge and level of precision the investigator would like to have about a particular factor ^74^.

Usability tests and interviews were most commonly used for involving users and capturing their perspectives across three stages of the medical device lifecycle. During usability tests, various data collection methods were used to collect both qualitative and quantitative data. Results such as time to complete tasks, or errors made during tasks, were easy to understand and compare. In its simplest form, a usability test just involved users performing a number of typical tasks and then reporting their experiences of using the device, i.e., what, in their opinion, worked well and what was problematic in the device placement ^43^. When testing an existing or prototype device to identify areas for improvement for example, qualitative data was more useful ^47^. In some of the reviewed studies, the developers observed users performing scenario-based usability tests to identify shortcomings or areas for improvement ^49^ and/or asked the user to report their experiences in follow-up interviews ^56^ or through user-centered methods such as empathy interviews, empathy mapping, and persona development as they complete the task ^64^. Sometimes quantitative data were collected during usability tests, such as the time taken to complete a task using the device or the number of errors made whilst performing the task ^42^. Interviews were mainly semi-structured and face-to-face. The factors on which information was routinely collected in these studies include socio-demographic characteristics, lifestyle practices, medical history, and use of medical devices.

Finally, our systematic review revealed that almost half of the papers we retrieved incorporated 2 or more methods to evaluate user perspectives throughout the MDD process. For example, a scoping method such as exploratory interviews ^65^ or a focus group ^51^ were used to specify the needs and requirements of the users with an evaluative method such as a usability test ^65^ or survey ^51^ then used respectively at a later stage to determine whether these have been met.

## 4. DISCUSSION

Successful medical device innovation requires research beyond technology-centred or technocentric design approaches to embrace user-centred methods 9. Employing formal methods to involve users in the MDD process should increase the probability that devices meet proper clinical goals, comply with technical standards, are cost-effective and meet ethical norms ^75^.

### 4.1 Types of devices

In this review a substantial proportion of the devices pertained to seizure detection, and healthcare and clinical information systems. This distribution aligns with previous studies ^77, 78^ that highlight the predominant focus on seizure forecasting and detection in epilepsy research, underscoring the profound impact of seizures’ unpredictability in this condition. Notably, only 5 VNS and 3 EEG based systems were found in our literature search. Moreover, among the 23 commercially available seizure detection devices and tools in the US and Europe, reported by Shum, Friedman ^79^, only Nightwatch, Epicare, Epilog, Sensor Dots and GeneActiv were identified to have involved users in the MDD process predominantly in the deployment phase ^27, 37, 39, 50^. This indicates a notable discrepancy in the extent of user involvement in the development of commercially available devices for epilepsy management. As highlighted by Hagedorn, Krishnamurty, Grosse ^80^, the disclosure of user involvement might be limited due to safety, privacy, and other ethical concerns that are unique to medical environments and contexts. Nevertheless, these results highlight a potential area for further research and improvement.

### 4.2 Types of users

The development of better products requires an in-depth understanding of all types of users, their activities, and their needs ^81^. In epilepsy, prior research has demonstrated that distinctions among users, including PWE, CG or HP can influence the delineation of user-specific requirements ^28^. Consequently, similar factors were appropriately considered in this study. Our review reveals that various types of medical devices are developed and assessed by PWE, CG and HP. These different groups of healthcare technology users’ characteristics, skills and working environment may be different, which is worth considering when developing health care technologies from the users’ perspective. It must be mentioned, however, that in assessing user needs, there is a substantial bias towards considering the needs of HP in contrast to PWE and CG. It is vital, therefore, that a range of users are consulted to get as wide an array of input as possible ^82^. Our review showed that PWE and CG are the users more involved in the MDD process. These users are likely to be a non-medical, heterogeneous group, including family members. As a result, their background, age, level of training, physical and mental fitness, and language knowledge vary considerably. Furthermore, the context of use is hard to predict and likely to be less controlled, when compared to a healthcare setting where standardised procedures and protocols are usually in effect. Therefore, understanding their individual needs is imperative for a successful device development process, and product quality and safety ^14, 16^, which might be the reason why these are more involved.

### 4.3 Methods of involvement

In the process of designing and developing medical devices, biomedical innovators have at their disposal a range of formal methods to incorporate user requirements. Our review shows that four methods were used to involve users in the MDD process, despite several other available approaches could have been considered such as heuristic evaluation, journey mapping^83^, and cognitive walkthrough ^84^. The majority of the studies used surveys, probably due to their effectiveness in gathering user feedback, namely through scalability, cost-effectiveness, standardization of data, anonymity, and quick data collection ^85^. With increasing attention being paid to patient-reported outcomes by funding agencies, measures of patient-centred factors, such as quality of life, depression, anxiety, cognitive, and functional status, are increasingly included in these surveys. Usability evaluation was the second most used method, reported in 20% of the reviewed studies, probably due to the current focus on improving usability and reducing human error within the medical device industry, and their need for regulatory agencies’ certification ^86^.

Depending on the purpose and context of the medical device, understanding user needs and eliciting their perspectives for developing medical devices could entail a combination of different methods ^14^. Our systematic review revealed that nearly half of the papers we retrieved incorporated two or more methods to evaluate user perspectives throughout the MDD process. The identified use of multiple qualitative and quantitative methods in combination with specific design methods is in line with the most recent literature which highlights the importance of multi-method human-centred design approaches for the development of health innovations comprising several design cycles ^87^.

### 4.4 Stages of MDD

Human-centred design should be central to all MDD to ensure that user needs are met. It is recommended that user-centred design should begin early, and continue throughout device development ^88^. Although we have found that users were involved in four out of the five stages of the MDD process - concept, design, testing and trials, and deployment stages - none of the medical devices identified was assessed in all of these four stages. Moreover the predominance of studies found in the deployment phase suggests that devices may lack sufficient user input during their development, since the form and function has already been determined, and the ability to innovate based on user needs is limited due to a number of fixed parameters ^3^. Consequently, the medical devices found in this review may only partially meet user requirements or fail to meet user requirements which could result in suboptimal user experiences, higher modification costs, and missed opportunities for innovation.

### 4.5 Regulatory view

Despite the fact that regulatory authorities such as Food and Drug Administration (FDA) and European Medicines Agencies (EMA) and international standards organizations have increasingly emphasized the importance of the employing formal methods for users’ involvement in the MDD as a critical factor in ensuring the effectiveness, safety, and usability of medical devices ^89, 90^, the involvement of users in the MDD process in epilepsy remains marginal. Our systematic review revealed that users were only involved in the MDD process of 13 commercially available medical devices for epilepsy management with most of the involvement being performed at the end of the MDD process in the deployment phase. These results are in line with other therapeutic areas, as medical device manufacturers often do not consider the benefit of employing formal human factors engineering methods within the MDD process^7^. This gap suggests that the appropriate employment of formal methods by manufacturers is unlikely to occur to significant levels without deliberate efforts to encourage and support manufacturers in doing so. Alternatively, the implementation of such methods could assume a mandatory character, being dictated to manufacturers by standards and purchasing agencies.

## 5. LIMITATIONS

This systematic review adopted the MDD cycle comprising 5 phases accordingly with Shah, Robinson ^16^. However, there is a lack of standardization of the MDD life cycle phases. WHO has yet to publish the medical device innovation part of the WHO medical device technical series, where it is expected to contribute to the standardization of the stages of the MDD process. Moreover, one limitation of this systematic review lies in the potential exclusion of medical devices developed by HP, whose contributions, though integral to the development phase, may not be explicitly reported in dedicated studies. Moreover, our review is contingent on published studies, and thus, it might not capture unpublished research, proprietary assessments by companies, or start-up initiatives aimed at gathering end-users’ feedback. This limitation highlights the potential underrepresentation of critical insights and developments in the field. The underreporting of user involvement in the MDD process, especially in epilepsy management, results in a missed opportunity for the broader medical community. Theoretically, once IP rights are secured, companies could safely disclose their methods of user involvement without jeopardizing their competitive position. However, the prevalent medical device industry practice of non-disclosure prevents the sharing of potentially beneficial experiences. Such transparency could foster an environment of collaborative improvement, motivating others to adopt user-centric approaches in device development. The lack of published information on user involvement methods in peer-reviewed journals hinders the opportunity for cross-learning and iterative enhancements in medical device design. This gap in knowledge dissemination ultimately impacts the quality and efficacy of devices developed for epilepsy care, as well as other medical fields. Additionally, our review found only 55 papers on user involvement or acceptability, likely because these metrics are often embedded within the paper rather than highlighted in the abstract or title. This may have inadvertently excluded relevant studies, suggesting there could be more instances of user involvement not captured in our review.

## 6. CLINICAL RELEVANCE OR FUTURE DIRECTIONS

This article provides a review and a conceptual understanding of the main user involvement research trends in MDD. The reviewed studies examined the medical-device usability concept from three viewpoints: PWE, CGs and HPs. In this review, 55 scientific sources related to user involvement in the medical device design and development in epilepsy were analysed and systematized. The following were discussed: (i) PWE are the users more involved in the MDD process; (ii) actual assessments may be applied differently for different medical devices with a higher prevalence for surveys and usability testing; (iii) formal methods for user involvement in the MDD process can be selected differently depending on the type of product; (iv) users are involved in 4 stages of the MDD process; (v) although regulatory guidelines are published to address user involvement in MDD process, future formal guidelines are needed to promote widespread of user involvement in MDD process in epilepsy.

## Data Availability

This is a systematic review

## Author Contributions

J.F., R.P., C.C., J.C. and L.L. were responsible for conceptualisation, study design, and search strategy for this Review. J.F. and R.P. carried out the systematic review. C.C. adjudicated disagreements. J.F. carried out the statistical analyses. J.F. and R.R. drafted the manuscript. All authors critically revised the manuscript, approved its submitted version, and accept responsibility for its content. The article is the original work of the authors and has not been previously published, nor is it currently under consideration by any other journal.

## Acknowledgments

This systematic review was funded by the European Union. Views and opinions expressed are however those of the author(s) only and do not necessarily reflect those of the European Union or European Innovation Council and SMEs Executive Agency (EISMEA). Neither the European Union nor the granting authority can be held responsible for them. The work of J.C. is partially financed by National Funds through the Portuguese funding agency, FCT - Fundação para a Ciência e a Tecnologia, within project LA/P/0063/2020. DOI 10.54499/LA/P/0063/2020 | https://doi.org/10.54499/LA/P/0063/2020. C.C. is supported by a Scientific Employment Stimulus contract (2020.00067.CEECIND) from Fundação para a Ciência e a Tecnologia.

## Ethical Approval

We confirm that we have read the Journal’s position on issues involved in ethical publication and affirm that this report is consistent with those guidelines.

## Appendix 1 – Data Extraction Form

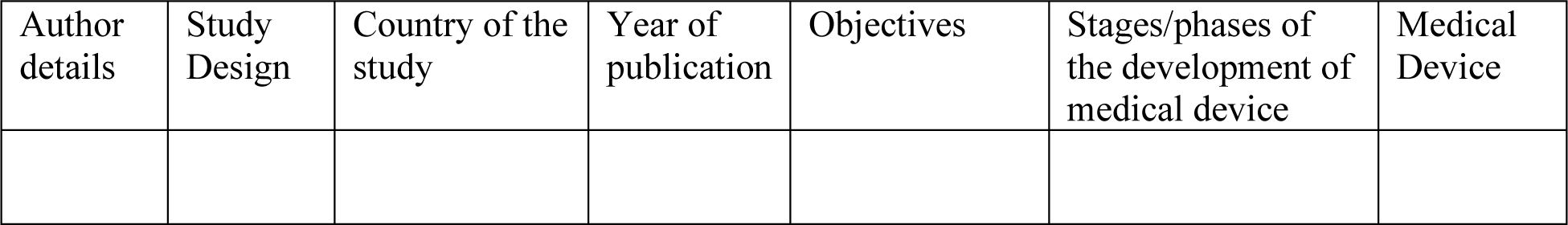

