## Supplemental Table S1 for "User involvement in the design and development of medical devices in Epilepsy: a systematic review"

**Supplementary Table S1**

Table 3 – Stages of MDD, an overview of the findings from the selected research studies

| Study, year of publication, country of origin | Objective of the study | Stages/phases of the development of the medical device | Methods used for capturing user perspectives by stages of the medical device lifecycle |
|---|---|---|---|
| Hufford, Glueckauf, Webb [17], 1999, US | To examine differences between the perceptions of adolescents with epilepsy and their parents in regard to comfort, distraction, and therapeutic alliance across 3 different modalities: (a) home-based video-system counseling, (b) home-based speakerphone counseling, and (c) videotaped, office-based counseling. | Deployment | Counseling sessions were conducted // After the assessment interview, each family member separately completed the Issue Severity Scale (ISS), Issue Frequency Scale (IFS), and Priority for Intervention Form of Glueckauf's Family and Disability Assessment System // clients completed the Audiovisual Equipment Rating Scale and the Audiovisual Equipment User Survey and a modified version of the Bond subscale of the Working Alliance Inventory for use in family counseling. They also completed the IPS, ISS, and Issue Change Scale (ICS) after the sixth session. |
| Zamponi, Rychlicki, Corpaci [18], 2008, Italy | To evaluate the safety and efficacy of vagus nerve stimulation (VNS) in very young children suffering from catastrophic epilepsy and status epilepticus. | Deployment | Quality of life was assessed using the Vineland Adaptive Behavior Scale (VABS) and by an analogical scale of parental satisfaction. |

| | | | |
|---|---|---|---|
| DiIorio, Escoffery, McCarty [19], 2009, US | To evaluate WebEase, an Internetbased,<br>theory-driven, self-management program<br>for adults with epilepsy. | Concept | Eligible and interested individuals were scheduled to attend one of three orientation sessions. At the beginning of the session, the participants completed an online pre-test about medication taking, stress and sleep. Following the survey, study staff presented an introduction to the WebEase Internet site. The participants were able to ask questions and test different components of the program using their own computer during orientation. |
| Schulze-Bonhage, Sales, Wagner [20], 2010, Germany, Portugal | To report on a two-center survey of patients´ views on the practical implementation of seizure prediction devices | Concept | 41 outpatients were asked to complete a survey designed to assess their views on the role of unpredictability of seizures, seizure prediction, requirements for the performance of prediction algorithms, and practical aspects of implementation |
| Koo, Yang, Seok [21], 2012, South Korea | To characterize epilepsy patients who use ‘Epilia’ as a main source of information compared to those who do not use ‘Epilia,’ but instead rely on healthcare providers for epilepsy-related information. The impact of using ‘Epilia’ on satisfaction and | Deployment | Separate surveys were administered to the online and offline groups to investigate their socio-demographic, epilepsy-related, and psychological characteristics, as well as |

| | | | |
|---|---|---|---|
| | attitude toward epilepsy was also investigated. | | attitude alterations after beginning to use 'Epilia'. |
| Vonhofen, Evangelista, Lordeon [22], 2012, US | To discuss the implementation of an automated injection system for isotope administration and its impact on staffing, safety, and nursing satisfaction. | Testing & Trials | The postimplementation satisfaction survey was sent to a cohort of 15 staff nurses |
| Thygesen, Sabers [23], 2013, Denmark | To evaluate the effect of VNS on seizure<br>frequency and to investigate patient satisfaction of and quality of life effects of VNS treatment. | Deployment | The study was an investigator-initiated retrospective survey of patients operated with VNS implantation during a ten-year period. |
| Brunnhuber, Amin, Nguyen [24], 2014, UK | To describe the development and implementation of video EEG telemetry (VT) in the patient's home (home video telemetry, HVT) in a single centre. | Testing & Trials | We used surveys to survey the principal stakeholders. consultants and technicians), to explore how HVT was perceived. |
| Askamp, van Putten [25], 2014, The Netherlands | We surveyed Dutch neurologists and patients and evaluated a novel mobile EEG device (Mobita, TMSi). | Deployment | Members of the Dutch Neurological Society were invited to participate in our online survey about the use of EEG in (suspected) epilepsy patients. |
| Hoppe, Feldmann, Blachut [26], 2015, Germany | To explore attitudes and preferences towards future devices for seizure detection in adult patients with therapy-refractory epilepsies. | Concept | 30-minute semi-structured interview on automated seizure registration |
| van Andel, Leijten, van Delden [27], 2015, Denmark | To describe stakeholders and their moral values that are relevant to the development and use of the seizure detector and present four design choices and the corresponding risks | Testing & Trials | Consortium Meetings, telephone conferences, email communications, survey |

| | | | |
|---|---|---|---|
| | and benefits in terms of the identified values. | | |
| Van de Vel, Smets, Wouters [28], 2016, Netherlands | To compare user experiences with systems for seizure detection, their opinions on usefulness and purpose of seizure detection, and their requirements for such a device. | Deployment | 2 Validated surveys: one for patients and caregivers and another for medical doctors |
| Newman, Shankar, Hanna [29], 2016, UK | To develop and implement an eHealth solution to support education and self-management of risks, in epilepsy. | Design, Test & Trials | Usability testing: user journeys, user observations, feedback, outcomes, app content evaluation |
| Patel, Moss, Rust [30], 2016, US | To inquire on detection device features that are important to patients and their caregivers. | Concept | A survey instrument |
| Sauro, Holroyd-Leduc, Wiebe [31], 2016, Canada | To develop a web-based, evidence-informed clinical decision tool to help physicians determine which patients are appropriate for an epilepsy surgery evaluation. | Deployment | Usability testing, focus groups, semi-structured interviews |
| Tovar Quiroga, Britton, Wirrell [32], 2016, US | To assess persons with epilepsy (PWE) and caregivers' perspectives regarding the features and priorities that should be considered in the design of seizure detection devices | Concept | Survey |
| Modi, Schmidt, Smith [33], 2017, US | to develop an individually tailored intervention, called Epilepsy Journey, to improve aspects of executive functioning through an iterative, patient-centered process including focus groups and usability testing. | Design | The unique needs of adolescents with epilepsy and their families were assessed based on focus groups. In the second phase, the web-based Epilepsy Journey problem-solving intervention was designed, developed, and evaluated. Evaluation took the |

| | | | |
|---|---|---|---|
| | | | form of multi-modal usability testing |
| Ali, Elsayed, Kaur [34], 2017, US | In a novel approach, we used social media to contact patients with DS to gather data on post-surgical seizure reduction and overall satisfaction with VNS | Deployment | A survey consisting of 10 questions was posted to a social media webpage for a DS support group moderated by the Dravet Syndrome Foundation. The results were analyzed, and percentages reported using the integrated SurveyMonkey analytical software. |
| Halford, Sperling, Nair [35], 2017, US | To evaluate the performance and tolerability in the epilepsy monitoring unit (EMU) of an investigational wearable surface electromyographic (sEMG) monitoring system for the detection of generalized tonic–clonic seizures (GTCSs). | Testing and Trials | A device usability survey was given to subjects. |
| Ozanne, Johansson, Hallgren Graneheim [36] 2018, Sweden | To explore perceptions regarding the use of wearable technology in disease monitoring and management as reported by individuals with epilepsy. | Deployment | Focus Groups and semi-structured interviews |
| Arends, Thijs, Gutter [37], 2018, Netherlands | To develop and prospectively evaluate a method of epileptic seizure detection combining heart rate and movement. | Testing & Trials | Survey (caregiver) and usability testing (PWE) |
| Afra, Bruggers, Sweney [38], 2018, US | To evaluate the patient's interest in using a mobile app for seizure control and self-care. | Concept | Survey |

| | | | |
|---|---|---|---|
| Meritam, Ryvlin, Beniczky [39], 2018, Denmark | To assess the performance, applicability, and usability of the device in home environment of patients | Deployment | Post-Study System Usability Survey (PSSUQ), telephone interviews |
| Janse, Dumanis, Huwig [40], 2019, US | To promote user-centered design of future seizure forecasting devices by quantifying patient and caregiver preferences for the potential benefits and risks of seizure forecasting devices. | Concept | Best-worst scaling (BWS) survey method |
| Chiang, Moss, Patel [41], 2020, US | To evaluate the association between seizure detection device usage and HR-QOL in an enriched population of SeizureTracker.com electronic seizure diary users. | Deployment | cross-sectional survey |
| Bruno, Biondi, Thorpe [42], 2020, UK | To assess the self-mastery performance and associated factors in a group of PWE admitted to an epilepsy monitoring unit who agreed to wear a wrist-worn device aimed at seizure detection | Deployment | Usability tests of the wearable device + Wearable technology Self-management Score (WSS) |
| Bruno, Biondi, Bottcher [43], 2020, UK, Germany | To identify the device placement-related issues experienced by a cohort of people with epilepsy (PWE) to investigate to what extent available devices are nonintrusive, comfortable, and stable on the body. | Deployment | Usability tests of the wearable device + Participants' experience was assessed using the Technology Acceptance Model Fast Form (TAM-FF) plus two additional questions on comfort. A thematic analysis was also performed on the free text of the survey. |
| Khan, Peechatka, Dias [44], 2020, US | To assess patient perceptions of completing ePROs, expectations of ePRO devices for PCOR and on-site | Concept | Online survey |

| | | | |
|---|---|---|---|
| | clinical visit in order to guide the development of successful ePRO deployment in seizure-related disorders. | | |
| Beck, Simony, Zibrandtsen [45], 2020, Denmark | To provide insights into patient readiness to use wearables for home monitoring of epilepsy | Deployment | Semi-structured qualitative interview 8 PWEs. |
| Thompson, Goodwin, Ojeda [46], 2020, US | Gain the perspectives of transition-age youth with epilepsy (TAYWE), their caregivers, and clinicians to inform the design of a mobile health (mHealth) system to support the self-management needs of TAYWE. | Deployment | Individual semi-structured interviews (Clinicians) and focus groups (PWE and caregivers) |
| Simblett, Matcham, Curtis [47], 2020, UK | To conduct a consultation exercise on the clinical end point or outcome measurement priorities for Remote Measurement Technologies studies, drawing on the experiences of people with chronic health conditions. | Concept | Focus Groups |
| Simblett, Biondi, Bruno [48], 2020, UK | Assess the first-hand experiences of people with epilepsy using wearable devices, continuously over a period of time. Understand how acceptable and easy they were to use, and whether it is reasonable to expect that people will use them. | Deployment | Semi structured interviews, Usability Test. |
| Yoo, Lim, Baek [49], 2020, South Korea | To develop a mobile epilepsy management application covering crucial factors comprehensively in a user-friendly way. | Design | Scenario-based usability test, satisfaction survey, and interview. |
| Nasseri, Nurse, Glasstetter [50], 2020, UK, US, | The present study addresses both of these problems of measuring device signal quality and assessing the | Deployment | At the end of the recording period, patients were provided with a survey to assess their |

| | | | |
|---|---|---|---|
| Germany, Australia | preferences of people with epilepsy while using wearable devices. | | preferences and comfort with using wearable devices for seizure prediction. |
| Buchhalter, Scantlebury, D'Alfonso [51], 2021, Canada | Describe the development of the Pediatric Epilepsy Outcome-Informatics Project (PEOIP) at Alberta Children's Hospital (ACH) | Testing and Trials | Meetings (HP), Survey (HP, CG) |
| Grzeskowiak, Dumanis [52], 2021, US | To understand the use-cases that best suit the needs of epilepsy community as seizure forecast research advances. These results will provide researchers with insight into user-acceptance of using a forecasting tool and incorporation into their daily life. Ultimately, this input from people living with epilepsy and caregivers will provide timely feedback on what the community needs are and ensure researchers and companies first and foremost consider these needs in seizure forecasting tools/product development. | Concept | A seizure forecasting survey targeted to those living with epilepsy and their caregivers was generated using Qualtrics (Provo, Utah) and distributed online |
| Cote, Beaudet, Auger [53], 2021, Canada | To evaluate the extent of utilization and acceptability of a web-based intervention among PWE | Testing and Trials | Pilot parallel-group randomized controlled trial 75 PWE who had Internet access allocated on a 1:1 ratio into an experimental group that received the intervention (experimental group (EG), n = 37) and a control group invited to consult epilepsy-related websites (control group (CG), n = 38). |

| | | | |
|---|---|---|---|
| Bruno, Biondi, Richardson [54], 2021, UK | To capture focal onset seizures with motor manifestations with a multimodal wearable device to identify the digital semiology and the evolution pattern of ictal manifestations. | Testing and Trials | Participants were asked to wear a multimodal wearable device (IMEC) aimed at seizure detection while admitted to an epilepsy monitoring unit. Seizures were labelled by a neurologist and start, and offset time were noted. The signals captured by the device during the seizure window were plotted and a visual inspection was performed for focal motor seizures with impaired awareness and for focal motor aware seizures |
| Herrera-Fortin, Bou Assi, Gagnon [55], 2021, Canada | To detail the preferences, needs and concerns regarding potential seizure detectors, of PWE and their caregivers across Canada. | Concept | Two online surveys were designed to survey PWE and their caregivers on seizure detection acceptability and to collect general clinical characteristics. |
| Olsen, Nielsen, Simony [56], 2021, Denmark | To explore the experiences of people with epilepsy using wearables for home seizure monitoring. | Deployment | Usability tests for 3 wearable devices, and semi structured open-ended individual interviews before and after home monitoring with wearable seizure monitoring equipment. |
| Willems, Baier, Bien [57], 2021, Germany | Satisfaction with and reliability of in-hospital video-EEG monitoring systems in epilepsy diagnosis - A German multicenter experience. | Deployment | Survey using a survey to assess the reliability, customer satisfaction, and potential for general or specific |

| | | | |
|---|---|---|---|
| | | | improvements of Video-electroencephalography monitoring systems |
| Monfort, Poulet, Nahas [58], 2021, France | This exploratory study aims to identify patient and caregiver groups, according to acceptability factors. | Concept | The 31-item Quality of Life in Epilepsy (QOLIE-31) scale, adapted from the more comprehensive 89-item scale; the 10 items Modified Computer Self Efficacy Scale (MCSES). The perception of the occurrence of seizures was evaluated using a Likert scale that ranged from 0 (never) to 5 (once or several times a day, every day).<br>A 21-item survey derived from the UTAUT 2 model was used to determine the connected patch acceptability. |
| Chiang, Moss, Black [59], 2021, US | Investigate patient and physician perspectives on effective translation of algorithm outputs into data visualizations through health information technology. | Design | We surveyed 627 people living with epilepsy and caregivers, and 28 epilepsy healthcare providers. Respondents scored each visualization in terms of international standardized software quality criteria for functionality, appropriateness, and usability. |
| Sabers, Aumuller-Wagner, Christensen [60], 2021, Denmark, | To explore potential clinical benefits of ta-VNS and to evaluate adaptation, compliance, as well as the usability of the device from a service design perspective. | Deployment | Service Design Survey on Medical Devices Used to determine if there were lifestyle and usability issues |

| | | | |
|---|---|---|---|
| Norway, the Netherlands, Belgium | | | |
| Siegel, Johnson, Dawson [61], 2021, US | To develop clinical decision support to reduce inappropriate prescription of carbohydrate-containing medications in hospitalized children on ketogenic diet. | Testing and Trials | Usability testing recommended by the National Institute of Standards and Technology: formative and summative testing |
| von Wrede, Rings, Schach [62], 2021, Germany | We developed an examination schedule to probe for immediate taVNS-induced modifications of large-scale epileptic brain networks and accompanying changes of cognition and behaviour. | Deployment | Survey on the evaluation of the device. Seven ordinal questions were asked concerning handling, possibility to continue activities while using the device, feeling while using the device, comfort, suitability for long-term and repeated use. |
| Hadady, Klivenyi, Fabo [63], 2022, Hungary, Denmark | Evaluate direct user experience with wearable seizure detection devices in the home environment. | Deployment | Structured online survey (175 caregivers and 67 persons with epilepsy) |
| Schmidt, Glaser, Riedy [64], 2022, US | To describe the iterative design, development, and evaluation of a novel mHealth learning environment for parents of children with epilepsy. | Design | Preparation included developing user personas and conducting focus groups. Phase 1 used empathy interviews, empathy mapping, and persona development methods. Usability and user experience methods were used, including SME reviews, cognitive walkthroughs, and usability testing. Survey |

| | | | |
|---|---|---|---|
| | | | responses and usability feedback were used to further refine the intervention. |
| Lazaro, Alvaran, Yun [65], 2022, South Korea | To develop a mHealth application for seizure management based on the human system integration (HSI) approach. | Concept, Design, Test & Trials | In the first phase, the needs were identified by conducting exploratory interviews with all the relevant stakeholders via e-mail or messaging app. In phase 2, to determine the system requirements, use case scenarios were employed wherein narratives or storyboards are created. As a supplement, personas and journey maps were also created to represent target users and their experiences. In the second phase, to determine the system requirements, use case scenarios were employed. In phase 3 a series of prototypes were created and tested. Users were also instructed to download and test the SeeSure app. The proposed app's function was evaluated through a focused-group interview involving the concerned stakeholders (1 PWE, 2 primary caregivers, 1 medical doctor) |
| Macea, Bhagubai, | To report the performance of an electroencephalogram (EEG) seizure- | Deployment | Patients used a WD, the Sensor Dot (SD), to measure two |

| | | | |
|---|---|---|---|
| Broux [66], 2023, Belgium | detector algorithm on data obtained with a wearable device (WD) in patients with focal refractory epilepsy and their experience. | | channels of EEG using dry electrode patches during pre-surgical evaluation and at home for up to eight months. An automated seizure detection algorithm flagged EEG regions with possible seizures, which we reviewed to evaluate the algorithm's diagnostic yield. In addition, we collected data on usability, side effects and patient satisfaction with an electronic seizure diary application (Helpilepsy®). |
| van Leeuwen, Droger, Thijs [67], 2023, Netherlands | To explore the perspectives of those receiving secondary care on nocturnal SDDs and epilepsy in general. | Concept | Interviews and Surveys |
| Guerrero-Aranda, Friman-Guillen, González-Garrido [68], 2023, Mexico | To assess the end-users acceptability of this proposed EEG report format. | Deployment | A 16-item electronic survey was sent to physicians who use EEG services of a medical diagnosis clinic. |
| Lewis-Fung, Tchao, Gray [69], 2023, Canada | Design and evaluate the feasibility of VR exposure therapy to treat epilepsy-specific interictal anxiety. | Design, Testing & Trials | online and in-person focus groups and survey |
| Tchao, Lewis-Fung, Gray [70], 2023, Canada | To explore and validate scenarios that provoke epilepsy/seizure-specific (ES) interictal anxiety and provide recommendations that lay the foundation for designing VR-ET | Concept | The survey included open- and closed-ended questions intentionally created to collect information useful for |

| | | | |
|---|---|---|---|
| | scenarios to treat this condition in PwE. | | informing the design of VR-ET scenarios for PwE |
| Lennard, Newman, McLean [71], 2023, UK | To evaluate the clinical utility and acceptability of Event Labs seizure monitoring (Nelli) in people with intellectual disability and epilepsy. | Deployment | A survey was developed by the clinical team in discussion. with experts by experience identifying preemptively what the key areas of inquiry need to be on the impact of Nelli. |
